# Decoding the genetic crosstalk network for cardiometabolic diseases and metabolic traits

**DOI:** 10.64898/2026.01.16.26344249

**Authors:** Aijie Li, Shufen Zheng, Zhuoyi Wu, Hanshuo Lu, Liang Chen, Cuiping Pan

## Abstract

Metabolic malfunctions are commonly observed in cardiometabolic diseases (CMDs), yet their genetic connections are not fully explored. We leverage multi-omics to decode the genetic crosstalk between 17 cardiometabolic diseases and 16 metabolic traits. Through genomic structural equation modeling, we identify 7 disease clusters anchored on 7 distinct metabolic axes, revealing subtype-specific variants, candidate genes, pathways, tissues and cell types. Among these are four metabolically distinct subtypes of arterial disorders, two adiposity subtypes with opposing associations with CMDs, and a heart-brain-kidney subtype characterized by blood pressure and BMI with dysregulated muscle-vessel coupling. Additionally, we uncover the genetic links between cardiometabolic processes and female health diagnoses. Finally, we leverage the shared risk genes to discover candidate drugs for CMDs, offering potential to improve comorbidity treatments. Overall, our study reveals the genetic basis of the metabolic networks underlying CMDs and extends their relevance into the female health. These results provide a foundation for future mechanistic studies and precise stratification of complex human diseases.

## Introduction

Cardiometabolic diseases (CMDs) encompass a range of cardiovascular and metabolic disorders that pose major challenges to global health^1^. Decades of epidemiological research have revealed that metabolic dysfunctions, such as hypertension, dyslipidemia, obesity, and impaired glycemic control, are key risk factors for the development and progression of CMDs^2^. Pioneering studies, including the landmark Framingham Heart Study, have shown that metabolic abnormalities significantly increased the risk of cardiovascular diseases^3,4^, and metabolic factors are effective for predicting CMD risks^5^. Thus, an inter-connected network exits among CMDs and metabolic traits.

Strong genetic foundations exit for CMDs and metabolic traits, with heritability estimates ranging from 40–70% for type 2 diabetes (T2D)^6^, 40–60% for coronary artery disease (CAD)^7^, 30-60% for hypertension (HTN)^8^, 40-80% for body mass index (BMI)^9^, 19-56% for waist-to-hip ratio (WHR)^10^, and 11-66% for blood lipid traits^11^. These substantial heritability values suggest that genetics plays a crucial role in shaping the relationships between CMDs and metabolic traits. However, existing studies examining the genetic crosstalk between CMDs and metabolic traits have largely focused on a limited number of phenotypes^3,4,12–21^, lacking a comprehensive view. Although family-based genetic correlation studies have demonstrated positive familial (co-)aggregation of cardiometabolic disorders^22^, in-depth exploration of biological mechanisms is lacking. Consequently, despite tremendous biological insights, contradictions have been observed in clinical practices. For example, while intensive lipid-lowering therapies have been shown to reduce ischemic stroke, they also increase the risk of intracerebral hemorrhage (ICH)^23^. Furthermore, glucose-lowering interventions have not consistently reduced cardiovascular disease (CVD) events^24^, and statin use has been linked to reduced CVD risks but increased incidence of T2D^25^. These contradictions emphasize the need to delineate the intricate inter-connections among CMDs and metabolic traits.

To overcome these challenges, we constructed a comprehensive network of genetic crosstalk among 17 CMDs spanning the major organ systems and 16 metabolic traits across the primary metabolic categories. Our analytical framework integrated genomic structural equation modeling (Genomic SEM)^26–28^, cross-trait meta-analysis, Mendelian randomization (MR) inference, variant annotation, biological pathway enrichment, and tissue/cell-type specificity analysis (**Fig. 1**). This approach enabled us to identify CMD subtypes characterized by distinct metabolic profiles, providing molecular insights into CMD classification and nominating candidate genes for the development of targeted therapeutics.

**Fig 1.**
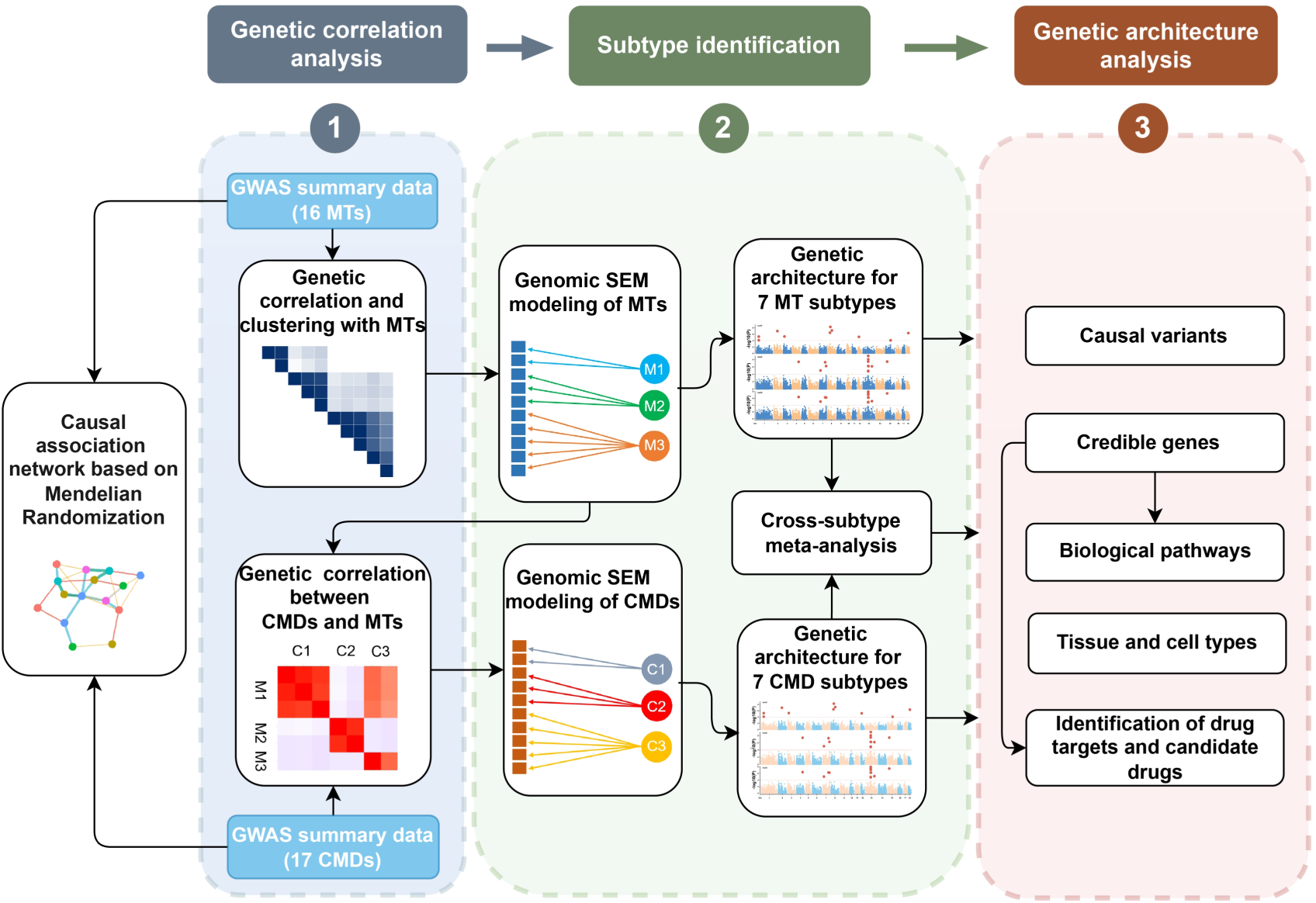
Study overview. Classification of cardiometabolic diseases (CMDs) and metabolic traits by genomic SEM, with identification of trait-specific and shared SNVs, genes, pathways, tissues, cell types, drugs and construction of causal inference networks.

## Results

### GWAS datasets and BMI adjustment

Our analysis incorporated comprehensive genome-wide association studies (GWAS) of 17 CMDs and 16 metabolic traits. The CMDs span five major biological systems: cerebrovascular, cardiovascular, circulatory, metabolic and endocrine, and renal systems. Meanwhile, the metabolic traits cover four metabolic categories: glycaemic, blood lipids, blood pressure, and adiposity-related traits (**Fig. 2a**). These GWAS summary statistics represent state-of-the-art genetic studies of cardiometabolism, with most datasets comprising more than 10,000 cases. The abundant significant SNVs (*p* < 5 × 10^−8^) provided substantial statistical power and robust signal for downstream analyses.

**Fig 2.**
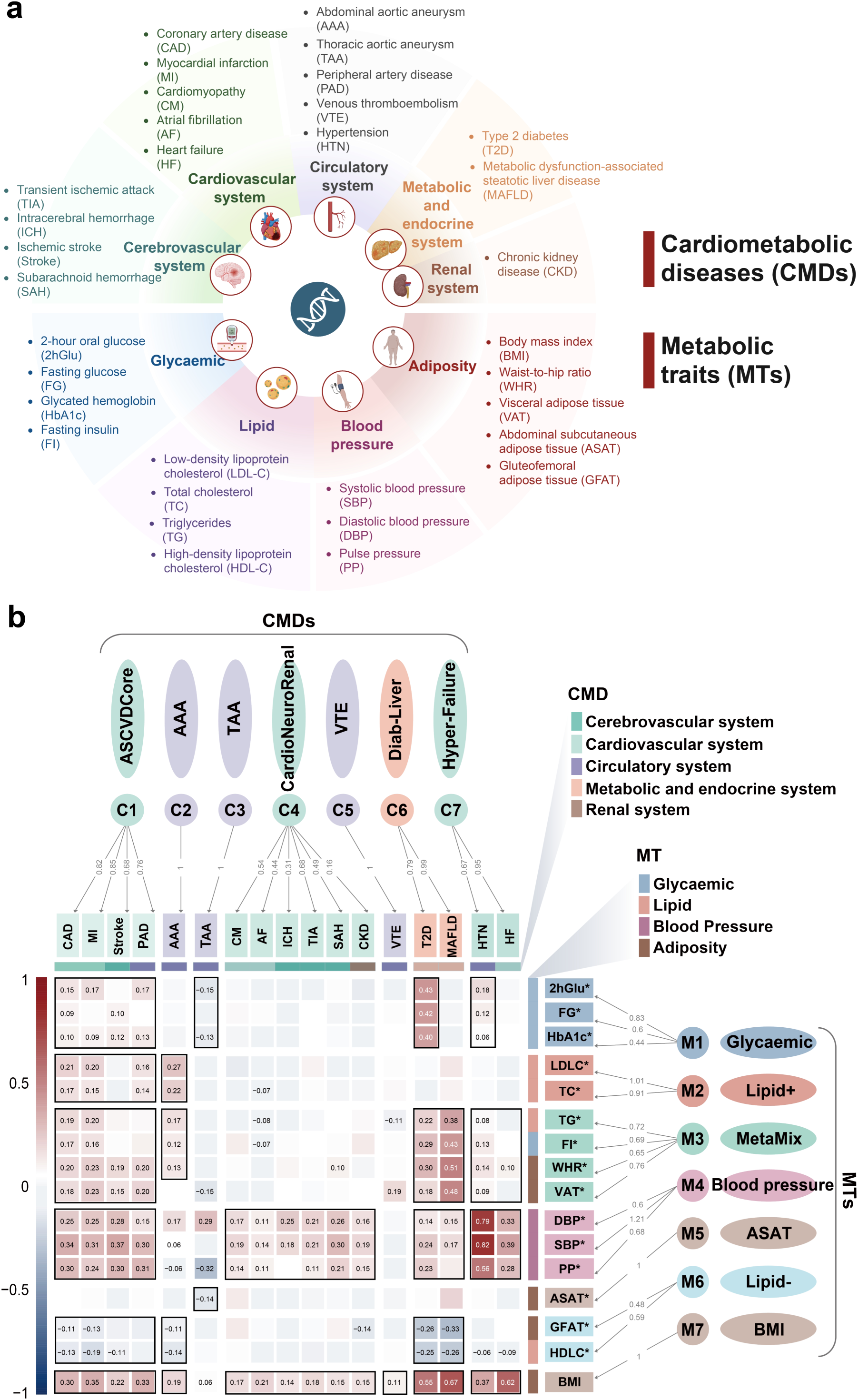
Genetic correlation between cardiometabolic diseases and metabolic traits. **a**. 17 cardiometabolic diseases (CMDs) and 16 metabolic traits (MTs) included in this study along with their clinical classifications (Created in BioRender. Pan, C. (2025) https://BioRender.com/4lkrdg2). **b**. Matrix of genetic correlation across seven CMD subtypes and seven MT subtypes. Color bars on the top and right indicate clinical classifications. Only the correlations (rg) passing statistically significant tests (FDR < 0.05) are displayed. Red and blue colors represent positive and negative genetic correlations, respectively. Color gradients indicate correlation strength. Genomic SEM-derived classifications are marked by black boxes. On the right, the arrows and numbers indicate standard loadings (one-headed arrows). CMD subtypes are denoted as C1 – C7 and MT subtypes as M1 – M7. As all GWAS datasets for MTs were adjusted for BMI, they are marked with an asterisk (*).

BMI was suggested to broadly influence metabolic phenotypes^14,29^, which we verified through comparing genetic correlations and causal relationships with or without considering BMI as covariates. Remarkably, the association of gluteofemoral adipose tissue (GFAT) with other traits was reversed after adjusting for BMI, rendering it consistent with epidemiological evidence^30^. Furthermore, BMI adjustment corrected several strong causal effects of metabolic perturbations on CMDs, e.g., the impact of blood pressure traits on thoracic aortic aneurysm (TAA). Meanwhile, this adjustment uncovered new causal relationships that would have otherwise been missed, including the effects of triglycerides (TG) and high-density lipoprotein cholesterol (HDL-C) on T2D and HTN. To minimize confounding effects, we used BMI-adjusted GWAS summary results for all subsequent analyses.

### Subtyping metabolic traits and CMDs

We examined the genetic correlations between CMDs and metabolic traits. Numerous CMDs, including CAD, myocardial infarction (MI), T2D, and metabolic dysfunction-associated steatotic liver disease (MAFLD), exhibited broad correlations with various metabolic traits. Conversely, several CMDs were primarily correlated with BMI and blood pressure traits, namely cardiomyopathy (CM), intracerebral hemorrhage (ICH), transient ischemic attack (TIA), subarachnoid hemorrhage (SAH), and chronic kidney disease (CKD). Specifically, venous thromboembolism (VTE) was exclusively associated with BMI.

These association profiles suggest genetic clustering patterns which may be modeled through Genomic SEM^26–28^. Indeed, seven metabolic clusters (M1-M7) were identified, achieving both excellent model fitting (comparative fix index (CFI) = 0.99, standard root mean square residual (SRMR) = 0.07) and highly representative subtype loci (*Q*_SNP_ heterogeneity tests). Integrating these metabolic subtypes into the genetic correlation matrix with CMDs, seven CMD clusters (C1-C7) with distinct metabolic correlation profiles were derived. We confirmed through Genomic SEM modeling that these seven CMD subtypes adequately described the genetic architecture of CMDs (CFI = 0.93, SRMR = 0.09; **Fig. 2b**), and most subtype loci were specific and representative of the subtype (*Q*_SNP_ heterogeneity tests).

Some of these genetics-driven metabolic subtypes, such as the glycaemic traits (M1) and blood pressure traits (M4), align with conventional clinical classifications that are typically guided by epidemiological and functional phenotypes. However, some metabolic subtypes present distinctions. For example, five obesity traits, including GFAT, visceral adipose tissue (VAT), abdominal subcutaneous adipose tissue (ASAT), WHR, and BMI, were clustered into four subtypes. GFAT clustered with HDL-C (M6), reflecting their common cardioprotective feature^31,32^. Although both visceral and abdominal subcutaneous fat were considered harmful^33^, VAT clustered with fasting insulin (FI), TG, and WHR in the M3 subtype, whereas ASAT formed an independent M5 subtype. BMI also formed an independent M7 subtype. Moreover, in contrast to its conventional association with glucose control, FI correlated more strongly with obesity traits (M3) than with glycaemic traits (M1). To reflect their specific characteristics, we named the subtypes as M1-Glycaemic (2-hour oral glucose [2hGlu], fasting glucose [FG], glycated hemoglobin [HbA1c]), M2-Lipid^+^ (low-density lipoprotein cholesterol [LDL-C], total cholesterol [TC]), M3-MetaMix (FI, TG, WHR, VAT), M4-Lipid^−^ (GFAT, HDL-C), M5-Blood Pressure (diastolic blood pressure [DBP], systolic blood pressure [SBP], pulse pressure [PP]), M6-ASAT, and M7-BMI.

Separately, the genetic subtypes of CMDs are largely concordant with clinical classifications^34–36^. The four atherosclerotic cardiovascular diseases (ASCVD), namely CAD, MI, stroke, and peripheral artery disease (PAD), co-clustered into C1 subtype and exhibited broad metabolic associations. T2D and MAFLD co-clustered into C6 subtype and displayed strong correlation with MetaMix (M3), Lipid^−^ (M4), and BMI (M7), but did not correlate with LDL-C or TC (M2). HTN clustered with heart failure (HF) into C7 subtype and showed the strongest genetic correlations with blood pressure traits among all CMD subtypes. These genetic profiles also reflect subtle differences. Although abdominal aortic aneurysm (AAA) was traditionally considered as ASCVD, it was set apart from other atherosclerotic diseases and formed an independent C2 subtype, due to its weak associations with glycaemic and blood pressure traits. Clearly, the two types of aortic aneurysms, TAA and AAA, have very different metabolic predisposition profiles. While AAA was impacted by blood lipids (M2) and adiposity traits (M3, M6, M7), TAA was inversely correlated by glycaemic traits (M1) and ASAT (M5). Notably, we identified the C4 subtype CardioNeuroRenal that encompassed diseases of heart (CM, atrial fibrillation [AF]), brain (ICH, TIA, SAH), and kidneys (CKD), which was exclusively characterized by blood pressures and BMI. In summary, these correlation patterns capture the major metabolic characteristics of CMD subtypes. As such, we defined these subtypes as C1-ASCVDCore (CAD, MI, stroke, PAD), C2-AAA, C3-TAA, C4-CardioNeuroRenal (CM, AF, ICH, TIA, SAH, CKD), C5-VTE, C6-DiabLiver (T2D, MAFLD), and C7-HyperFailure (HTN, HF).

Collectively, our analysis resulted in seven metabolic subtypes and seven CMD subtypes. Among the 49 possible subtype pairs, 24 exhibited significant genetic correlations. Summarizing their correlation strengths, we found that ASCVDCore (C1) was impacted by 6 metabolic subtypes, followed by DiabLiver (C6), HyperFailure (C7), and AAA (C2), which were impacted by 5, 4, and 4 metabolic subtypes, respectively. TAA (C3), VTE (C5), and CardioNeuroRenal (C4) subtypes showed selective associations with only one or two metabolic subtypes. In summary, distinct metabolic characteristics underlie different CMD subtypes.

### Coherent causal relationships across CMDs and metabolic traits

Genetic associations do not necessarily imply causal effects. To infer causality, we performed bidirectional MR analyses across the CMDs and metabolic traits. A total of 235 (22.2%) causal relationships were identified, including 47 from CMDs to other CMDs, 95 from metabolic traits to CMDs, 23 from CMDs to metabolic traits, and 70 from metabolic traits to other metabolic traits. Based on these, we constructed networks for the positive (**Fig. 3a**) and the negative causal relationships (**Fig. 3b**).

**Fig 3.**
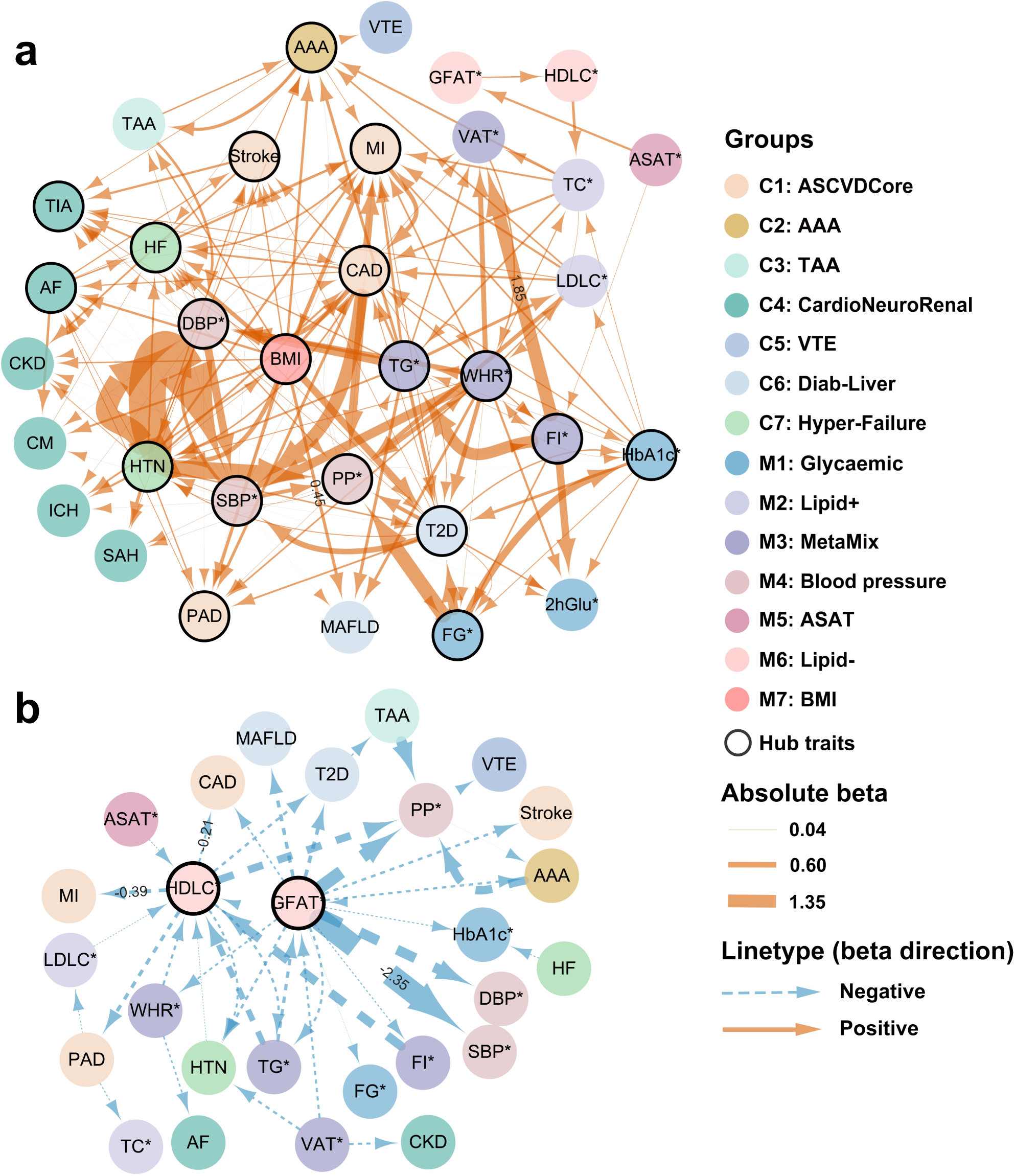
Causal linkage network between CMDs and metabolic traits. The bidirectional causal relationship across four categories, i.e., MT→CMD, CMD→CMD, MT→MT, and CMD→MT, are illustrated for (**a**) positive causality and (**b**) negative causality. Nodes represent traits, colored according to seven CMD groups (C1-C7) and seven MT groups (M1-M7). Directed edges (arrows) indicate causal effects inferred by MR. Orange solid lines (β > 0) and blue dashed lines (β < 0) denote effect directionality, with edge thickness scaled to absolute β-values (see scale bar). Networks were pruned to show only statistically significant associations (FDR <0.05). Nodes with black border are hubs (with association links ≥ 9) in the network.

BMI (M7) and blood pressure traits (M4), consistent with their broad genetic correlations, exerted broadest positive causal effects to CMDs, with BMI causal to 15 CMDs, DBP causal to 15 CMDs, SBP causal to 14 CMDs, and PP causal to 13 CMDs. Conversely, GFAT and HDL-C in the Lipid^−^ subtype (M6) exerted broad protective effects. Other notable hubs are the MetaMix subtype (M3: FI, TG, WHR, VAT), being causally impacted by 14 metabolic traits and in its downstream, exerting causal impacts on 24 traits, which in total accounted for 67% (32/48) of the positive causal linkages within the metabolic traits. Therein, TG and WHR were causal to 7 and 6 diseases, respectively. CardioNeuroRenal (C4) is driven solely by blood pressures and BMI. These results largely mirror the genetic correlation results, supporting the validity of the subtype classification. Notably, we found that the HTN-CAD-T2D module forms a fully bidirectional causal loop, accounting for 62% (29/47) of the causal linkages within CMDs, suggesting their central roles in CMD co-occurrences.

### Genetic architecture of the subtypes

We then decoded the genetic architecture for all 14 subtypes through SNVs, genes, pathways, tissues, and cell types. Significantly associated SNVs can be extracted from the latent factors after the Genomic SEM modeling. These SNVs were seldom shared across subtypes, validating the specificity of the modeling. Through fine mapping, we identified 8,736 causal variants (PIP > 0.9) for the CMD subtypes, which were annotated to 551 credible genes by three complementary approaches. Comparing with literature, 24% of the genes have been uniformly reported in all diseases within a CMD subtype, validating the effectiveness of our method. Additionally, 36% of the genes have been previously associated with a single disease but are now suggested by our study as relevant to other diseases within the subtype. The remaining 40% of the genes have not been associated with any disease within the subtype. Similar to SNVs, these genes exhibited strong subtype specificity, with only 76 genes (14%) shared across subtypes, such as *LPL* and *SLC22A3* shared by ASCVDcore (C1), AAA (C2), and DiabLiver (C6). Mutational hotspots for cardio-metabolism were revealed, with the most striking examples as *CDKN2B-AS1* on chr9, shared in ASCVDcore (C1), AAA (C2), and DiabLiver (C6), as well as *CFDP1* on chr16, shared in ASCVDcore (C1), AAA (C2), and HyperFailure (C7) (**Fig. 4a**). Furthermore, the subtype genes were enriched for pathways that aligned with their correlated metabolic patterns (**Fig. 4b**). ASCVDcore and AAA converged on pathways of plasma lipoprotein particles and cholesterol metabolism. DiabLiver was enriched for hormone secretion, pancreatic islet, and glucose response. HyperFailure was enriched for blood pressure regulation and kidney functions. These findings are further corroborated by studies investigating the pathogenesis of CMD comorbidity^34–36^. Notably, CardioNeuroRenal (C4) was characterized by cardiac muscle contraction and muscle tissue development.

**Fig. 4.**
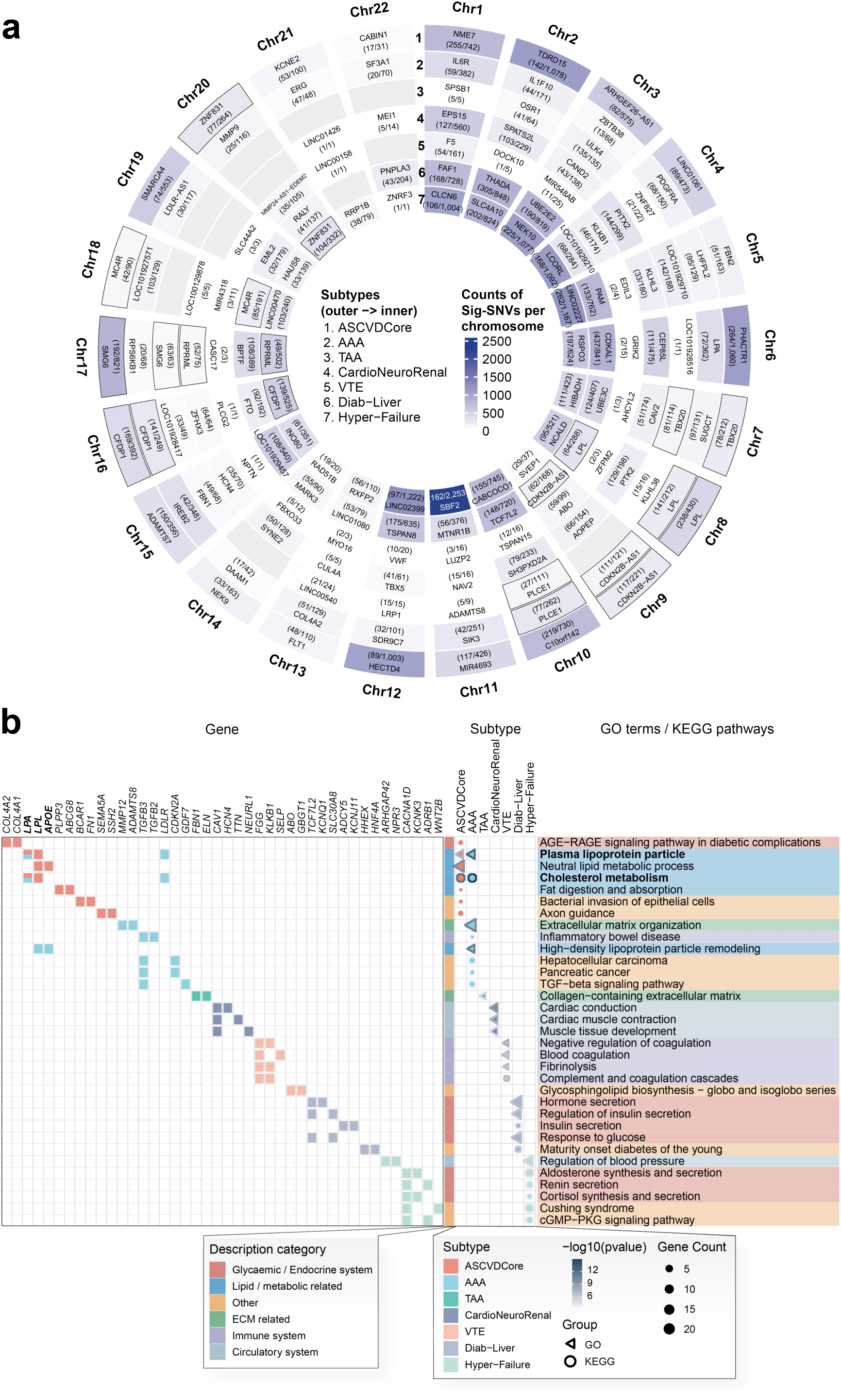
Molecular features of CMD subtypes. **a.** Representative genes by chromosome for each subtype. Circular plot shows the per-chromosome counts of genome-wide significant SNVs (p < 5e-8) for each subtype across the 22 autosomes. A white-to-steelblue gradient denotes the counts; autosomes with no significant SNVs was marked with light gray. The outer ring labels chromosomes (“Chr”), and rings from outside to inside represent subtypes (C1-ASCVDCore, C2-AAA, C3-TAA, C4-CardioNeuroRenal, C5-VTE, C6-DiabLiver, C7-HyperFailure). For each subtype, each cell marks the gene with the highest number of significant SNVs, and reports the number of significant SNVs (p < 5e-8) mapped to this gene relative to the total number of significant SNVs on the corresponding chromosome. Cells with black borders have repetitive representative genes across different subtypes. **b.** Pathway enrichment for subtype-specific genes. Triangles represent GO terms and circles represent KEGG terms. Symbol size indicates gene count, and outer color intensity reflects –log₁₀ (p) values. The left side displays the two genes with the strongest effect for each term. Repeated terms or genes are bolded, and different groups are indicated by distinct colors.

We also analyzed the metabolic subtypes (M1-M7) and identified 36,127 causal variants and 1,975 credible genes. 349 genes (18%) were shared by multiple subtypes, with the most pleiotropic genes such as *PPARG, GCKR, TCF7L2, GRB14,* and *COBLL1* found in at least four metabolic subtypes. These genes have been known to play versatile roles in regulating glucose, lipid, and metabolic processes. Interestingly, in addition to its established associations with glucose and lipid metabolism, the MetaMix subtype (M3) was significantly enriched for immune-related processes, suggesting a link between metabolic regulation and immune function. The enriched pathways reflect known functional specificity of each subtype^37–39^. For example, genes in the glycaemic subtype (M1) were primarily enriched for glucose regulation and insulin secretion, genes in the blood pressure subtype (M4) were enriched for kidney and muscle-related functions, and genes in the BMI subtype (M7) were enriched for brain and neural pathways.

### Genetic crosstalk between the CMD subtypes and metabolic subtypes

Given the strong sharing of genetic basis between CMDs and metabolism, we analyzed the crosstalk between 24 subtype pairs. A total of 1,744 credible genes and 286 causal loci were identified after meta-analysis and colocalization. The enriched pathways for each subtype pair conform to its correlated metabolic subtypes, reflecting the strong metabolic predisposition underlying each CMD subtype.

We further identified the associated tissue and cell types through heritability enrichment. To integrate cells with tissues, we computed cell type compositions for each tissue leveraging the human body atlas of single-cell transcriptome and single-cell chromatin accessibility, followed by constructing the trait–cell–tissue network. Across the 14 subtypes and the 24 disease-trait subtype pairs, 58 enriched cell types spanning 16 tissues were identified (**Fig. 5a**). Mesenchymal and supporting cells are dominant, including fibroblasts, fetal mesangial cells, pericytes, and smooth muscle cells. The dominant tissues include vascular/cardiovascular compartments (artery, heart, blood vessels, muscle), metabolic/endocrine organs (pancreas, liver, adipose, adrenal gland, kidney), and the gastrointestinal tract. Together, vascular/stromal structure, tissue remodeling, and endocrine–metabolic processes are suggested essential to cardiometabolic regulation. Note that these tissue and cell enrichment patterns align with their corresponding pathway profiles. BMI (M7) and its subtype pairs were primarily enriched for brain tissues, while subtypes involving glycaemic traits (M1) were enriched for pancreas, gastrointestinal tract, and liver. Across all subtypes, ASCVDCore (C1), MetaMix (M3), and Blood Pressure (M4) were associated with more than 10 tissues and mediated by fibroblasts, pericytes and endothelial cells (**Fig. 5a**), which aligned with their broad impact in the causality networks, underscoring the complex interplay in these traits. Conversely, some subtypes exhibited specific tissue enrichment patterns: Lipid^−^ (M6) was selectively enriched for liver and adipose tissue (**Fig. 5c**), VTE (C5) was enriched for liver and heart, and HyperFailure (C7) was enriched specifically for the adrenal gland.

**Fig. 5.**
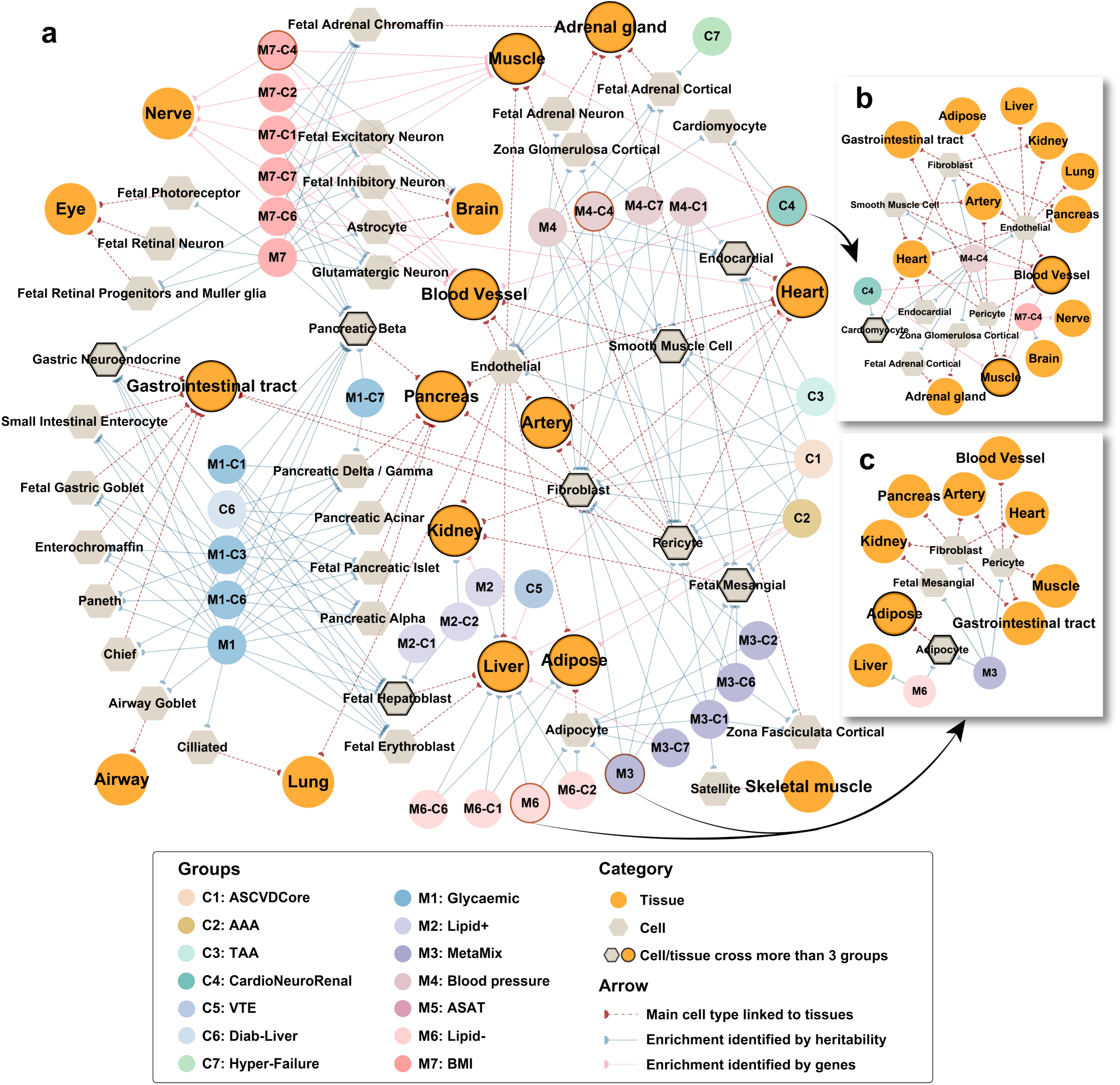
Tissue and cell type-specific network of cardiometabolic subtypes. **a.** The connections among heritability-enriched tissues and cell types across subtypes. **b.** The heritability-enriched tissue and cell-type network for the CardioNeuroRenal (C4) subtype. **c.** The heritability-enriched tissue and cell-type network for the MetaMix (M3) and Lipid- (M6) subtypes. Orange circles denote tissues; hexagons denote cell types. Colored circles mark the subtypes or subtype pairs, as shown in the legend. Blue lines mark the results from heritability enrichment analysis. Pink lines mark additional links derived from gene-level enrichment. Red dashed lines denote major cell type–tissue assignments inferred from single-cell datasets from healthy individuals. Black borders highlight tissues or cell types shared by three or more groups.

The shared architecture of the disease-trait subtype pairs has provided us rich genetic insights into distinct metabolic regulation under different CMDs. In the following, we highlight several discoveries.

#### (1) Stratification of arterial disorders

In our framework, diseases of the arterial system are distributed in four subtypes: ASCVDCore (C1: CAD, MI, stroke, PAD), AAA (C2), TAA (C3), and CardioNeuroRenal (C4: TIA) (**Fig. 2b**). We revealed a pan-metabolic dysregulation in ASCVDCore characterized by extensive links to glucose, blood pressure, lipids, and obesity traits, affecting all major cardiometabolic organs and primarily mediating through endothelial cells. Conversely, AAA exhibits a unique metabolic profile with selective dysregulation in lipids and adiposity. In addition, pronounced ECM dysregulation and inflammatory responses were identified for AAA, with smooth muscle cells and fibroblasts rather than endothelial cells as the primary cell types. Nonetheless, lipid metabolism remains a common predisposition feature for ASCVDcore and AAA. Among the 171 credible genes for ASCVDCore and the 92 genes for AAA, 27 genes were shared and most of them were associated with lipid metabolism, including *APOB, CETP, LIPA, LPA, LPL,* and *SCARB1*.

While AAA is a disorder of the abdominal section of the aortic aneurysm^40^, TAA is a disorder of the thoracic section^41^. Our analysis suggests that both AAA and TAA are enriched for smooth muscle cells, fibroblasts, and artery tissues. However, a thorough examination reveals that elastic fiber formation and ECM integrity, but not lipid-related processes, are strongly enriched for TAA. Thus, although AAA and TAA share similar histological features, their underlying genetic origins are distinct. TIA, a known outcome of ASCVD, was classified to the CardioNeuroRenal subtype (C4) as it was mostly tied to blood pressure and BMI.

#### (2) Cardiometabolic-protective genes

Two metabolic subtypes consistently exhibited opposing effects to other traits (**Fig. 3**). While MetaMix (M3: FI, TG, WHR, VAT) drives CMD risks, Lipid^−^ (M6: GFAT, HDL-C) exerts a protective effect. Both metabolic subtypes are significantly associated with ASCVDcore (C1), AAA (C2), and DiabLiver (C6). They also share cellular and tissue types, notably adipocytes, adipose tissue, and liver (**Fig. 5c**). Pathway enrichment confirmed their common involvement in lipid-related processes. However, while Lipid^−^ was exclusively lipid-focused, MetaMix engages additional non-lipid pathways, including diabetes and immunity. Scanning genes associated with both subtypes in the liver and adipose tissue may help locate cardiometabolic-protective genes. As such, we identified 42 genes associated positively with Lipid^−^ while negatively with MetaMix. These genes represent 25% of Lipid^−^ credible genes but only 12% of those in MetaMix. Among the list, the protective roles of *KLF14, COBLL1, SHBG,* and *LPL* have been experimentally validated in mouse or cellular models^42–45^.

#### (3) Cross-organ metabolic characteristics of the CardioNeuralRenal axis

We discovered a subtype CardioNeuroRenal (C4) that spans the cardiac (CM, AF), cerebral (ICH, TIA, SAH), and renal (CKD) functions. Our convergent genetic evidence indicates blood pressure and BMI as the primary metabolic susceptibility in this subtype, whereas lipid and glucose traits are not genetically associated with it. The heart–brain–kidney axis is an emerging observation in epidemiological studies, with heighten blood pressure as a risk factor^12,46,47^. We identified 58 credible genes for this subtype, among which *CAV1, KLHL3* and *FBN2* had been reported in some of these diseases. *CAV1* and *KLHL3* encode regulators of cardiovascular excitability and vascular tone, whereas *FBN2* is involved in maintaining the sarcomere and cytoskeletal integrity in myocytes. We also identified genes such as *KCND3*, *CAMK2D*, and *NR3C1*, which are closely related to cardiac muscle function and blood pressure regulation. Pathway and tissue/cell analysis converged on the heart, vasculature, muscle, smooth muscle cell and cardiomyocyte (**Fig. 4b and Fig. 5b**), implicating muscle contraction as a shared mechanism. Together, our analysis suggests hemodynamics and cardiac muscle regulation as a common genetic predisposition in the CardioNeuroRenal axis.

#### (4) Genetic crosstalk between cardiometabolic traits and female health diagnoses

Mammary gland and the female reproductive tract (e.g., uterus, cervix, and fallopian tube) were repeatedly enriched across multiple cardiometabolic subtypes including ASCVDCore (C1), AAA (C2), TAA (C3), MetaMix (M3), and Blood Pressure (M4), prompting us to investigate the genetic basis of cardiometabolic traits and female health diagnoses. We therefore collected GWAS studies of 43 female health diagnoses^48,49^, covering disorders of the female genital tract, pelvic organs, complications related to pregnancy or labor, and neoplasms, for analyzing their shared genetic architecture with the 14 cardiometabolic subtypes. Although these female health diagnoses studies had smaller sample sizes compared to the CMDs, they offered a preliminary overview.

We observed 126 pairs of significant associations among female health diagnoses and multiple cardiometabolic subtypes, with the former category including 25 conditions such as “edema, proteinuria and hypertensive disorders in pregnancy, childbirth and the puerperium”, “other maternal disorders predominantly related to pregnancy”, “inflammatory diseases of female pelvic organs”, “leiomyoma”, and “complications of labor and delivery”, and the latter category including ASCVDCore (C1), AAA (C2), CardioNeuroRenal (C4), DiabLiver (C6), HyperFailure (C7), MetaMix (M3), Blood Pressure (M4), and BMI (M7). The MetaMix subtype (M3) were broadly linked to 14 female health diagnoses, with shared genes exhibiting enriched heritability in breast tissue, adipose tissue, adipocytes, and fibroblasts. Additionally, uterus, cervix, and fibroblasts were enriched by the shared genes between 7 female health diagnoses and 5 CMD subtypes, confirming a common genetic basis between the cardiometabolic subtypes and the female health diagnoses.

Bidirectional causal inference suggested 48 causal relationships (β > 0.1) between the two disease categories, most of which (35/48, 73%) were cardiometabolic traits exerting a causal effect on female health diagnoses. BMI was inferred to causally raise the risks of 8 female health outcomes. In addition, we identified positive causal effects of AAA on “hydatidiform mole”, HTN on “excessive, frequent and irregular menstruation” and “other female pelvic inflammatory diseases”, and CM on “premature separation of placenta”. Reverse effects were also suggested, such as a positive causal effect of “female infertility associated with anovulation” on AAA and “leiomyoma of the uterus” on CM. These results suggest mutual influences between cardiometabolic traits and female health diagnoses.

We conducted Summary-data-based Mendelian Randomization (SMR) to identify the shared genes. A total of 1,310 genes were identified, which were mostly shared by female health diagnoses with MetaMix (M3), BMI (M7), HyperFailure (C7), DiabLiver (C6), and ASCVDCore (C1). Of these, 471 genes (36%) were identified in highly relevant tissues such as adipose tissue, breast tissue, fibroblasts, and uterus. Evidence of colocalization suggests 92 SNVs (PPH4 > 0.5) and 52 genes are shared between 11 cardiometabolic subtypes and 10 female health diagnoses. Among the list are genes involved in transcriptional regulation, such as *TCF7L2*, *MAFF*, *KLF14*, *HMGA1*, and *ZNF831*. Other key genes, shared by at least 4 disease pairs, include *FGF5*, *FTO*, *CDKAL1*, and *ARHGAP42*. Therein the fat mass and obesity-associated gene *FTO*, which encodes an N^6^-methyladenosine demethylase capable of reversing m6A, has been proven essential for adipogenesis^50^ and associated with female infertility and reproductive system diseases^51^, thus linking adiposity with female conditions. Considerable potential for dual effects may exit for the other identified genes in both cardiometabolic regulation and female conditions.

### Potential therapies for treating CMD comorbidities

Our delineation of the CMD subtypes reveals potential comorbidity basis, which can inform candidate therapies. To this end, we prioritized disease genes with consistent effects in all the primary tissues of each CMD subtype. Drugs considered were cardiovascular therapies, metabolic disease therapies, immunosuppressants, anti-inflammatory natural products, and other agents with potential cardiovascular benefits, of which the action directions were carefully examined. Following that, pairing between the pathological and pharmacological pathways^52^ was computed, resulting in 61 drugs (pathway matching score > 0.3) that targeted 25 risk genes, forming 71 drug–target-subtype associations that were distributed in 6 CMD subtypes (C1,C2, C4-C7) (**Fig. 6**). 42% (30/71) of these associations have existing clinical evidence (with regulatory approval or demonstrated efficacy in clinical trials) across all diseases within the same subtype. Examples include antihypertensive agents for HyperFailure (C7), such as Carvedilol (targeting *KCNH2* and *ADRB1*), Metoprolol (targeting *ADRB1*), and Quinapril (targeting *ACE*). Antidiabetic drugs were identified for DiabLiver (C6), including Dulaglutide (targeting *GLP1R*), Repaglinide, and Rosiglitazone (targeting *PPARG*).

**Figure 6.**
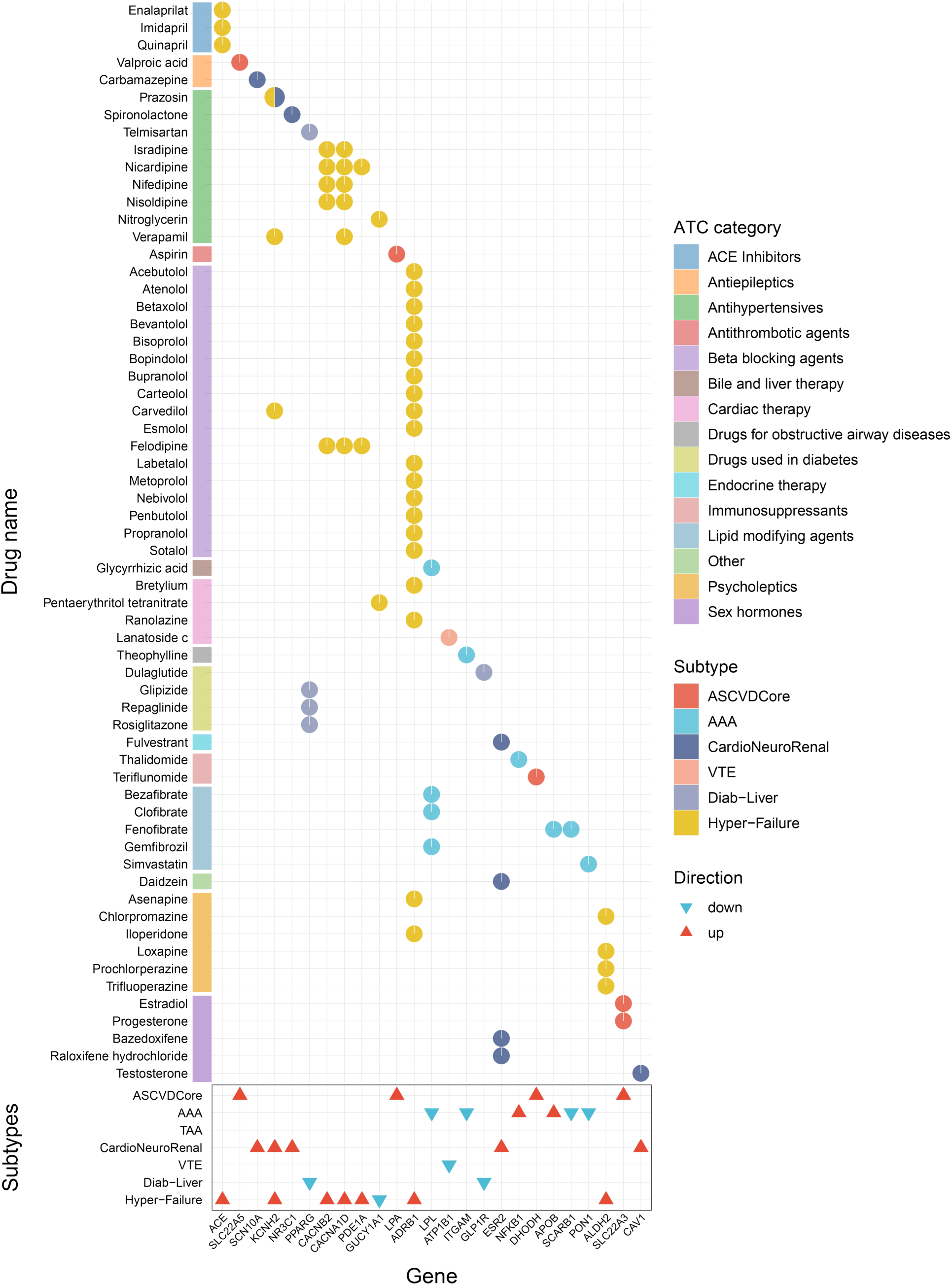
Potential drug targets and drugs associated with CMD subtypes. The lower box illustrates the association between CMD subtypes and genes, with triangular symbols indicate the association direction of the gene with the CMD subtypes. The upper section displays drugs that acting on the genes, with circular pie charts denoting the subtypes in which the drug-gene pair were identified. The color bars on the left indicate the Anatomical Therapeutic Chemical (ATC) classification of drugs.

Beyond recapitulating known therapies, our analysis also uncovered opportunities for drug repurposing in CMDs. We identified 41 previously underexplored drug-gene–subtype associations. An example is the immunosuppressant Teriflunomide (targets *DHODH*) for ASCVDcore (C1). While it is currently approved for treating relapsing–remitting multiple sclerosis^53^, animal studies suggest that Teriflunomide may mitigate atherosclerosis by modulating lipid metabolism and endothelial dysfunction^54^, thereby warranting further investigation of its treatment potential in ASCVD. Another example is the sex hormone Testosterone (targets *CAV1*) for CardioNeuroRenal (C4). The potential benefits of Testosterone has been reported in observational studies of AF^55^ and experimental models of SAH^56^, although the mechanisms remain unclear. In addition, several psycholeptics that primarily targets *ADRB1* and *ALDH2*, including Asenapine, Trifluoperazine, Iloperidone, and Loxapine, were identified as candidate drugs for HyperFailure (C7). Moreover, the AAA subtype was linked to lipid-lowering agents such as Simvastatin, which has been shown to attenuate aneurysm formation in mouse models^57^, although this has yet to be confirmed by clinical trials. We also identified additional candidates for AAA, including Theophylline (targets *ITGAM*) in treating obstructive airway diseases, Gemfibrozil (targets *LPL*) in lowering blood lipids, and the immunosuppressant Thalidomide (targets *NFKB1*), all of which may provide potential drug repurposing opportunities.

## Discussion

In this study, we identified seven CMD subtypes and seven metabolic subtypes through genetic architecture modeling, followed by depicting the distinct genetic features for each subtype and their crosstalk. This genetically informed taxonomy complements symptom-based classification, clarifies genetic mechanisms of CMD comorbidity, and provides a molecular framework for developing precise therapeutics.

A key finding is that distinct metabolic axes underlie each CMD subtype, which reinforces the notion that the genetic basis of CMDs is tightly coupled with metabolic dysregulation^2,22,58^, rendering our disease subtyping highly interpretable. While some genetic subtypes conform to clinical subtyping^37,39^, such as the glycaemic traits (M1: 2hGlu, FG, HbA1c)^39^ and the blood pressure traits (M4: DBP, SBP, PP)^37^, some are markedly different. We examined the five arterial disorders that spread in four CMD subtypes, various lipid and adiposity traits in Lipid^−^ (M6) and MetaMix (M3), and a novel CardioNeuroRenal subtype (C4) that spans a long axis of organ systems, concluding that strong and specific metabolic predisposition indeed underlie each subtype, which are consistently manifested in the levels of genes, biological pathways, tissues, and cell types.

Our efforts revealed both genetic homogeneity and heterogeneity across different subtypes, resulting in a complex genetic network. BMI represents a low-resolution obesity phenotype exhibiting broad, non-specific adverse effects on cardiovascular metabolic health^14,29^, highlighting the necessity of BMI correction during data integration, particularly for obesity-related traits. BMI and blood pressure exhibit broad genetic associations with CMDs. In addition, the metabolic traits of TG and WHR, as well as the diseases CAD, T2D, and HTN, were identified as hubs in the overall cardiometabolic processes. Thus, we provide supportive genetic evidence that further reinforces their central roles in cardiometabolic regulation^2,58,59^.

Lipid and adiposity traits are central to CMDs. We identified a metabolic subtype, MetaMix (M3), which displayed broad positive associations with CMDs including ASCVDCore (C1), AAA (C2), DiabLiver (C6), and HyperFailure (C7). Traits included in this subtype are visceral fat, triglycerides, fasting insulin, and waist-to-hip ratio. These traits form central hubs in the positive causality network among all metabolic traits. Besides, there are ample evidence in literature to report their strong association and important roles in CMDs^60,61^, suggesting their essential roles in cardiometabolic regulation. Separately, we discovered a cardiometabolic-protective subtype Lipid^−^, which included GFAT and HDL-C, corroborating the known protective roles of these two traits^31,32^. There seems to be a genetic equilibrium between MetaMix and Lipid^−^, with lipid metabolism as a shared pathway between the risky and protective conditions. Our study also clarified the genetic distinction between fat distributions. While fat distributed in subcutaneous abdomen (M5: ASAT) was classified as an independent subtype with negligible genetic predisposition to cardiometabolic health, visceral fat (M3: VAT) was identified as a harmful core for cardiometabolism. This finding is contradictory to the existing assumption that both fat types are detrimental^33,62,63^ and may contribute to an explanation for “metabolically healthy obesity”.

Our study further reveals genetic connections between cardiometabolic traits and female health outcomes. Earlier studies have revealed gender differences in CMDs^64^. Our analysis uncovers broad genetic sharing between cardiometabolic traits (C1: ASCVDCore, C7: HyperFailure, M3: MetaMix, and M6: BMI) and female health diagnoses, which contributes to understanding the metabolic drivers underlying female health outcomes and the gender differences in CMDs. Moreover, we also identified the shared causal SNVs, risk genes, and relevant cell and tissue types behind these genetic connections, providing potential directions for future research on the “metabolism–reproduction axis.”

Importantly, the genetic subtyping of CMDs offers the potential to leverage genetic information to advance therapeutic development. We identified 61 candidate drugs, with 42% of the drug–subtype associations supported by clinical approval or clinical trials. The alignment between the drug categories and the metabolic signatures across CMD subtypes confirmed the biological coherence of the inferred therapies. Moreover, the 41 underexplored drug–subtype associations offer opportunities for repurposing existing drugs for CMDs and expanding treatment options for comorbidity management. Understanding the metabolic predisposition profiles may inform precise treatment strategies. Particularly, knowing effect directions of the shared genes in multiple diseases or subtypes may help navigate drug side effects, so as to minimize therapeutic contradictions in clinical practices.

There are several limitations in our study. First, although we nominate putative CMD-protective genes, functional studies in animal or human models are required to interrogate their mechanisms. Second, GWAS of mostly European ancestry was investigated in this study. Due to insufficient traits available in other ancestral groups, multi-ancestry validation is challenging. The expanding GWAS in non-European population will enable assessment of the generalizability of our discoveries. Third, although a primary association between cardiometabolic traits and female health outcomes was identified, the limited sample sizes of the female health outcomes has limited our discovery power. Finally, we utilized reference datasets of healthy tissues to interpret gene regulation and expression for disease signals, which may neglect disease-specific signals. Future studies leveraging disease-specific single-cell transcriptomic, spatial transcriptomic, epigenomic and other data types will further refine our interpterion of the genetic basis for complex human diseases.

## Methods

### GWAS datasets

We obtained 47 GWAS summary statistics from public repositories and consortia, including Center for Statistical Genetics (CSG), NHGRI-EBI GWAS catalog, FinnGen Release 10, Common Metabolic Diseases Knowledge Portal, The MEGASTROKE consortium, Million Veterans Program (MVP), the DIAGRAM Consortium, the CKDGen Consortium Meta-Analysis Data, the Meta-Analyses of Glucose and Insulin-related traits Consortium (MAGIC) and GIANT consortium, covering 17 CMDs and 31 MTs (16 BMI-adjusted, 14 unadjusted). All summary statistics were harmonized to GRCh37 reference genome using easylift (v1.0.0), annotated with SumStatsRehab, and subjected to genotype quality control, excluding variants lacking rsIDs, variants with minor allele frequency (MAF) < 1%, and duplicate variants). SNP effects were standardized as Z-scores (Z = β/SE) for cross-study comparison.

### Genetic correlation estimates

Genetic correlations were estimated using linkage disequilibrium score regression (LDSC) v1.0.1^65^, which regresses GWAS χ^2^ statistics on SNP linkage disequilibrium (LD) scores to account for confounding due to population stratification and polygenicity. Summary statistics were harmonized to HapMap3 European reference via munge function, using LD scores from the 1000 Genomes European population. False discovery rate was controlled with the Benjamini-Hochberg method (FDR < 0.05). Partitioned heritability was performed using stratified LDSC (S-LDSC)^66^.

### Mendelian randomization

We performed bidirectional Mendelian randomization (MR) analyses for all included traits using five complementary MR methods, including inverse variance weighting (IVW)^67^, MR-Egger^68^, weighted median^69^, CAUSE^70^, and MR-APSS^71^. The IVW method served as the primary analysis, providing unbiased estimates under the assumption of balanced pleiotropy. MR-Egger regression was used to detect directional pleiotropy, where a significant intercept (*p* < 0.05) indicated a violation of the instrument strength independent of direct effect assumption. The weighted median method provided consistent estimates when up to 50% of the instrumental weight came from invalid instruments. CAUSE employed Bayesian modeling to distinguish between correlated and uncorrelated pleiotropy, while MR-APSS applied adaptive pruning to select optimal instrument subsets and adjust for sample overlap and pleiotropic pathways. To ensure accurate interpretation of MR estimates, we applied sensitivity analyses including heterogeneity testing (Cochran’s Q), pleiotropy assessment (MR-Egger intercept and MR-PRESSO), and leave-one-out analysis.

Genome-wide significant SNVs (*p* < 5 × 10^−8^) were selected as initial instrumental variables (IVs). LD clumping (r^2^ < 0.001, 1,000 kb) was performed using the 1000 Genomes European reference panel to ensure independence, implemented in PLINK (v1.9). Only variants with an F-statistic > 10 (F = (R^2^(N−2))/(1−R^2^)) were retained. For CAUSE and MR-APSS, the default thresholds of p = 1 × 10^−3^ and p = 5 × 10^−5^ were applied. Meanwhile, we excluded variants with known phenotypic associations (p < 1 × 10^−6^) using the GWAS Catalog through FastTraitR (v1.0.0). Credible causal relationships were defined by consistent effect direction across all methods, statistical significance (p < 0.05) in at least two methods including IVW, and no evidence of horizontal pleiotropy (MR-Egger intercept p ≥ 0.05). When potential pleiotropy was indicated by MR-Egger, MR-PRESSO^72^ was applied to validate and correct for horizontal pleiotropy. P-values were adjusted using FDR < 0.05. All analyses were performed using the R packages TwoSampleMR (v0.6.7)^73^, MR-APSS (v2.0.0), CAUSE (v1.2.0) and MR-PRESSO (v1.0). Genomic coordinates were standardized to GRCh37/hg19, and network graphs were visualized using Cytoscape (v3.10.3).

### Subtype model specification and GWAS estimation with Genomic SEM

#### Genetic correlation estimation and phenotypic clustering

We first estimated genetic correlations using the LDSC function in Genomic SEM^26–28^. GWAS summary statistics were filtered by the HapMap3 European panel and the LD reference panel obtained from the Genomic SEM repository. For MT grouping, we first modeled the conventional clinical classifications, but the residual fit was not ideal. We then performed exploratory factor analysis (EFA). Specifically, genetic correlations were clustered using hierarchical clustering, which revealed seven distinct clusters. To validate whether this classification adequately explained the genetic architecture of MTs, we calculated model fit indices using Genomic SEM, considering CFI > 0.95 and SRMR < 0.10 as indicators of good fit. The results confirmed the validity of the seven-cluster classification, which provided a better fit than the conventional clinical subtypes.

Next, we attempted to model the conventional clinical classifications of CMDs, but the model could not be successfully constructed. Moreover, given the extensive genetic correlations among CMDs, clustering based solely on CMD correlations was insufficient for clear delineation. To address this, we implemented a two-step strategy that integrated both CMD genetic associations and their genetic relationships with MTs. First, we performed primary clustering based on genetic correlation patterns aligned with established pathophysiology (e.g., ASCVD [CAD, MI, Stroke, PAD]; T2D and MAFLD; HTN and HF). In the second step, we refined the classification of ambiguous traits according to their genetic associations with the pre-defined MT subtypes. Final validation using Genomic SEM, after systematically testing models with one to nine factors, confirmed that the seven-CMD-subtype solution provided the best fit for CMD subtyping and, importantly, revealed distinct and robust genetic boundaries between CMDs and MTs.

#### Shared genetic architecture analysis

To characterize shared genetic architecture across different groups, we modeled the genetic variance-covariance matrices for each group. Shared SNP effects within groups were estimated using the weighted least squares (WLS) method with the userGWAS function. To evaluate whether the computed SNP effects operated through our classification model, we performed *Q*_SNP_ heterogeneity testing using independent SNPs. The heterogeneity test produced χ^2^ statistics, where the null hypothesis suggested that SNPs were functioning through the specified model (*p* > 5×10^−8^). Thus, rejection of the null hypothesis indicated that SNPs operated through a different model than the specified one.

### Cross-trait meta-analysis

To identify shared genetic architecture between CMD-MT subtype pairs, we integrated multi-trait analysis of GWAS (MTAG)^74^ and cross-phenotype association analysis (CPASSOC)^75^. MTAG jointly analyzed trait pairs under the assumptions of homogeneous SNP heritability and perfect genetic covariance, this approach leverages genetic correlations to improve power for detecting pleiotropic loci. CPASSOC was performed using the SHet statistic to robustly detect cross-trait associations despite effect size heterogeneity. SNVs achieving genome-wide significance in both methods (*P*_MTAG_ & *P*_CPASSOC_ ≤ 5×10^−8^) were retained, and independent loci were subsequently defined using LD clumping (r*^2^*<0.1, within a 1000 kb window).

### Fine-mapping analysis

We identified a 99% credible set of causal variants through a Bayesian fine-mapping method named FM-summary^76^. Independent index variants were identified with PLINK v1.9 (--clump-p1 5 × 10^−8^ --clump-r2 0.1 --clump-kb 1000), and variants within ± 500 kb were extracted as candidate regions. Variants with posterior inclusion probability (PIP) > 0.9 were considered high-confidence causal candidates.

### Gene annotation

We applied two methods to identify genetic predisposition genes. Gene-based association was performed through MAGMA (v1.10)^77^ and GCTA-fastBAT^78^. MAGMA aggregated SNP-level summary statistics across protein-coding gene regions (n = 18,719 genes) using gene-based test statistics. While GCTA-fastBAT performed gene-based association tests using a fast set-based approach that accounts for LD among SNPs. Both methods applied stringent multiple testing correction (Bonferroni-adjusted *p* < 0.05) to prioritize robust associations.

Transcriptome-wide association analysis was performed using Summary-data-based Mendelian Randomization (SMR)^79^ and FUSION^80^ methods. SMR analysis integrated GWAS summary statistics with cis-eQTL data from GTEx V8 (49 tissues, hg19). Using genome-wide significant SNPs (*p* < 5×10^−8^) as IVs, with HEIDI-outlier tests (*p* > 0.05, requiring ≥ 3 independent SNPs) distinguishing causal relationships from linkage. FUSION tested the aggregate effect of all cis-variants per gene using pre-computed expression weights from GTEx v8 data, analysis restricted to autosomal chromosomes (chr1 - 22) and excluded the major histocompatibility complex (MHC) region (chr6: 25 - 34 Mb) due to its complex LD structure. Both methods accounted for LD structure using 1000 Genomes Project European reference panels. Significant genes in both methods were defined as passing both Bonferroni-adjusted *p* < 0.05 (with tissue-specific correction applied).

### Credible gene sets identified

We defined fine-mapping–based causal genes as the nearest annotated genes to fine-mapped SNVs using ANNOVAR (hg19)^81^. Credible genes were defined as the overlap of results from three complementary approaches: gene-based association, transcriptome-wide association, and fine-mapping-based annotation.

### Colocalization

Colocalization analysis was performed to identify shared causal variants between subtype pairs. For each pair (CMD subtype–MT subtype; original GWAS–CMD subtype), SNVs within ± 500 kb of lead variants from subtype summary statistics were extracted. Posterior probabilities for five hypotheses were estimated using the coloc (v5.2.3)^82^ package in R, with PP.H4 representing the posterior probability of a shared causal variant. Loci with PP.H4 > 0.7 were considered to provide evidence of colocalization.

### Enrichment analysis

Gene Ontology (GO) and Kyoto Encyclopedia of Genes and Genomes (KEGG) pathway enrichment analyses were conducted on the gene set using the clusterProfiler (v4.12.0)^83^. GO enrichment was performed for the biological process (BP), molecular function (MF), and cellular component (CC). Significance was defined as Benjamini–Hochberg FDR < 0.05.

### Tissue and cell specific enrichment

Tissue and cell-specific enrichment analysis was performed using LDSC regression for specifically expressed genes (LDSC-SEG)^84^, which estimates the proportion of SNP heritability explained by tissue/cell-specific binary SNP annotations, with the resulting test statistics serving as evidence for tissue/cell relevance. We utilized pre-computed LD scores derived from tissue-specific gene expression data (53 tissues from GTEx) ^84^ and candidate cis-regulatory elements (cCREs) across 222 cell types from CATLAS^19,85^.

We further performed MAGMA^77^ to validate the tissue-specific results from LDSC-SEG, which aggregates SNP-level GWAS signals into gene-level associations and tests for enrichment in specific tissues. Notably, nearly all significant tissue enrichments identified by LDSC-SEG were validated by MAGMA (FDR < 0.05). Moreover, we implemented a gene annotation-based approach using the Tissue Specific Expression Analysis (TSEA) website^86^, which provided additional tissue enrichment results.

To identify linkage between tissues and cells, we calculated the predominant cellular composition of each major tissue using data from publicly available single-cell datasets from healthy individuals. The predominant cell type was determined by superimposing cell type proportions and applying a 95% threshold.

### Potential drug target identification

To systematically identify potential therapeutic targets, we conducted drug-target MR analyses. Putative genes were defined by integrating results from FUSION, SMR, GCTA-fastBAT, MAGMA, and fine-mapping analyses. These genes were mapped to eQTL-associated genes using the GTEx v8 dataset, which comprises 15,201 RNA-sequencing samples across 49 tissues from 838 postmortem donors^87^. Additionally, pQTLs from seven plasma proteome-wide association studies (covering 4,853 proteins)^88^ were used to assess the causal effects of plasma proteins on different CMD subtypes.

IVs were filtered to ensure validity: we included 1,851 cis-pQTLs and cis-eQTLs associated to putative genes (variants located within ± 1 Mb of the gene transcription start site), requiring F-statistics > 10 to avoid weak instrument bias, genome-wide significance (*p* < 5 × 10^−8^), and LD clumping (r^2^ < 0.001 within 1 Mb windows) to ensure independence. MR analyses were conducted using the Wald ratio (for single IVs) or IVW method (for multiple IVs), with statistical significance determined after FDR < 0.05.

To minimize potential side effects of on-target drug activity, only targets showing concordant effect directions in primary tissues (as defined by LDSC-SEG tissue and cell specific enrichment) across FUSION, SMR, MR, eQTL and pQTL analyses were retained.

### Candidate drug identification

We compiled potential drug targets for seven CMD subtypes and identified candidate therapeutic drugs using a five-step process. First, we collected drugs associated with these potential targets from eight gene–drug interaction databases (TTD, Pharos, PharmGKB, DrugBank, DrugCentral, DGIdb, CTD, and DSigDB). We then applied two filtering criteria: (i) drugs must be recorded in at least two databases as interacting with the target gene; (ii) drugs belonged to one of five predefined categories—cardiovascular therapies, metabolic disease therapies, immunosuppressants, anti-inflammatory natural products, or other agents with potential cardiovascular benefits. Drug action direction was further considered to ensure suitability for each CMD subtype. To compare enrichment profiles between pathological and pharmacological gene sets, we used a pathway pairing score method^52^. Pathological pathways were defined by enrichment of subtype-associated genes, while pharmacological pathways were based on enrichment of drug targets from above databases, with enrichment analysis performed using clusterProfiler (v4.12.0). Finally, subtype–gene–drug combinations with pathway matching scores greater than 0.3 were retained as candidate therapeutic associations.

## Data Availability

All data produced in the present study are available upon reasonable request to the authors

## Abbreviations

2hGlu: 2-hour oral glucose
AAA: Abdominal aortic aneurysm
AF: Atrial fibrillation
ASAT: Abdominal subcutaneous adipose tissue
ASCVD: Atherosclerotic cardiovascular disease
BMI: Body mass index
CAD: Coronary artery disease
CKD: Chronic kidney disease
CM: Cardiomyopathy
CMD: Cardiometabolic disease
CVD: Cardiovascular disease
DBP: Diastolic blood pressure
FG: Fasting glucose
FI: Fasting insulin
Genomic SEM: Genomic structural equation modeling
GFAT: Gluteofemoral adipose tissue
GWAS: Genome-wide association studies
HbA1c: Glycated hemoglobin
HDL-C: High-density lipoprotein cholesterol
HF: Heart failure
HTN: Hypertension
ICH: Intracerebral hemorrhage
LDL-C: Low-density lipoprotein cholesterol
MAFLD: Metabolic dysfunction-associated steatotic liver disease
MI: Myocardial infarction
MR: Mendelian randomization
MT: Metabolic trait
PAD: Peripheral artery disease
PP: Pulse pressure
SAH: Subarachnoid hemorrhage
SBP: Systolic blood pressure
T2D: Type 2 diabetes
TAA: Thoracic aortic aneurysm
TC: Total cholesterol
TG: Triglycerides
TIA: Transient ischemic attack
VAT: Visceral adipose tissue
VTE: Venous thromboembolism
WHR: Waist-to-hip ratio

## Data availability

The GWAS summary statistics used in this study are available from:

CSG (https://sph.umich.edu/csg/), NHGRI-EBI GWAS catalog (https://www.ebi.ac.uk/gwas/), FinnGen Release 10 (https://r10.risteys.finngen.fi/), Common Metabolic Diseases Knowledge Portal (https://hugeamp.org/), MEGASTROKE (http://megastroke.org/), MVP (https://www.research.va.gov/mvp/), DIAGRAM (https://www.diagram-consortium.org/), CKDGen (https://ckdgen.imbi.uni-freiburg.de/), MAGIC (https://magicinvestigators.org/) and GIANT (https://giant-consortium.web.broadinstitute.org/GIANT_consortium). Software support data were obtained from Genomic SEM LD reference panel (https://github.com/GenomicSEM/GenomicSEM/wiki), SMR cis-eQTL data (https://yanglab.westlake.edu.cn/data/SMR) and FUSION (http://gusevlab.org/projects/fusion), 1000 Genomes Project European reference panels (https://cncr.nl/research/magma/#reference_data), ANNOVAR (https://annovar.openbioinformatics.org/en/latest/user-guide/download) and GTEx eQTL (https://www.gtexportal.org/home/downloads/adult-gtex/qtl). The websites used in this study are TSEA (http://doughertytools.wustl.edu/TSEAtool.html), TTD (https://ttd.idrblab.cn/), Pharos (https://pharos.nih.gov/), PharmGKB (https://www.clinpgx.org/), DrugBank (https://go.drugbank.com), DrugCentral (https://drugcentral.org/), DGIdb (https://dgidb.org/), CTD (https://ctdbase.org/), and DSigDB (http://tanlab.ucdenver.edu/DSigDB).

## Code availability

Publicly available software (and version, where applicable) used in this paper is as follows: easylift (v1.0.0, https://github.com/zhanghaoyang0/easylift); SumStatsRehab (https://github.com/Kukuster/SumStatsRehab);; LDSC (V1.0.1, https://github.com/bulik/ldsc); TwoSampleMR (v0.6.7, https://github.com/MRCIEU/TwoSampleMR); CAUSE (v1.2.0, https://github.com/jean997/cause); MR-APSS (v2.0.0, https://github.com/YangLabHKUST/MR-APSS); MR-PRESSO (v1.0, https://github.com/rondolab/MR-PRESSO). PLINK (v1.9, https://www.coggenomics.org/plink2/); FastTraitR (v1.0.0, https://github.com/TullyMonster/MendelRookie); Cytoscape (v3.10.3, https://manual.cytoscape.org/en/stable/); Genomic SEM (https://github.com/GenomicSEM/GenomicSEM); Pheatmap (https://github.com/raivokolde/pheatmap); corplot (https://github.com/taiyun/corrplot); MTAG (https://github.com/JonJala/mtag); CPASSOC (http://hal.case.edu/~xxz10/zhu-web/CPASSOC); FM-summary (https://github.com/hailianghuang/FM-summary); MAGMA (v1.10, https://cncr.nl/research/magma/); GCTA-fastBAT (https://yanglab.westlake.edu.cn/software/gcta/#fastBAT); SMR (https://yanglab.westlake.edu.cn/software/smr); FUSION (http://gusevlab.org/projects/fusion/); ANNOVAR (https://github.com/WGLab/doc-ANNOVAR); coloc (v 5.2.3, https://github.com/chr1swallace/coloc); clusterProfiler (v4.12.1, http://bioconductor.org/packages/release/bioc/html/clusterProfiler.html); LDSC-SEG (https://github.com/bulik/ldsc). Other code is available at GitHub (https://github.com/aijieli1/The-Human-Health-Blueprint).

## Acknowledgments

This study was supported by the research grants awarded to C.P. from National Natural Science Foundation of China (No. 32270626) and National Key Research and Development Program of China (No. 2024YFC2707700). We thank members of the Laboratory of Intelligent Computing in The Hong Kong University of Science and Technology (Guangzhou) for insightful discussions and suggestions.

## Contributions

C.P. and A.L. conceived and designed the study. A.L. conducted data collection, analyses and result visualization. S.Z. performed supplementary drug analyses. Z.W. contributed additional analyses on female health diagnoses. A.L. wrote the original draft. C.P., L.C., Z.W., S.Z., and H.L. provided critical inputs for revising the manuscript. All authors agreed to the final version of the manuscript.

